# Methodological choices in brucellosis burden of disease assessments: A systematic review

**DOI:** 10.1101/2022.05.10.22274867

**Authors:** Carlotta Di Bari, Narmada Venkateswaran, Mieghan Bruce, Christina Fastl, Ben Huntington, Grace Patterson, Jonathan Rushton, Paul Torgerson, David Pigott, Brecht Devleesschauwer

## Abstract

**Background:** Foodborne and zoonotic diseases such as brucellosis present many challenges to public health and economic welfare. Increasingly, researchers and public health institutes use disability-adjusted life years (DALYs) to generate a comprehensive comparison of the population health impact of these conditions. DALYs calculations, however, entail a number of methodological choices and assumptions, with data gaps and uncertainties to accommodate. The following review identifies existing brucellosis burden studies and analyzes their methodological choices, assumptions, and uncertainties. The review supports the Global Burden of Animal Diseases programme in the development of a systematic methodology to describe the impact of animal diseases on society, including human health.

**Methods / Principal findings:** A systematic search for brucellosis burden calculations was conducted in pre-selected international and grey literature databases. Using a standardized reporting framework, we evaluated each estimate on a variety of key methodological assumptions necessary to compute a DALY. Thirteen studies satisfied the inclusion criteria (human brucellosis and quantification of DALYs). One study reported estimates at the global level, the rest were national or subnational assessments. Data regarding different methodological choices were extracted, including detailed assessments of the adopted disease models. Most studies retrieved brucellosis epidemiological data from administrative registries. Incidence data were often estimated on the basis of laboratory-confirmed tests. Not all studies included mortality estimates (Years of Life Lost) in their assessments due to lack of data or the assumption that brucellosis is not a fatal disease. Only two studies used a model with variable health states and corresponding disability weights. The rest used a simplified singular health state approach. Wide variation was seen in the duration chosen for brucellosis, ranging from 2 weeks to 4.5 years, irrespective of the whether a chronic state was included.

**Conclusion:** Available brucellosis burden assessments vary widely in their methodology and assumptions. Further research is needed to better characterize the clinical course of brucellosis and to estimate case-fatality rates. Additionally, reporting of methodological choices should be improved to enhance transparency and comparability of estimates. These steps will increase the value of these estimates for policy makers.

**Author Summary:** Brucellosis is a bacterial disease transmitted to humans by consumption of contaminated, unpasteurized milk or through direct contact with infected animals and their excretions. This disease causes production losses and has major economic impacts on individuals and communities. The disability-adjusted life year (DALY) is a metric for measuring the burden. It summarizes mortality (years of life lost) and morbidity (years lived with disability) into a single metric. This review aimed to identify existing brucellosis burden studies and analyse their methodological choices, assumptions, and uncertainties. The results suggested that some parameters carry considerable uncertainty, particularly mortality and disease duration. This highlights the importance of strengthening routine reporting systems, collecting better mortality data and conducting further research on the course of brucellosis. Additionally, estimates of DALYs will benefit from a deeper understanding of the symptoms and the different sources of attribution. Finally, current reporting of methodological choices should be improved to enhance transparency, comparability, and consistency of brucellosis burden.

## Introduction

Brucellosis is globally one of the most widespread zoonoses and is caused by the genus *Brucella* [1]. This bacterial disease can be transmitted from animal reservoirs, such as cattle, sheep, goats, and pigs, to humans, through consumption of raw animal products, most commonly unpasteurized milk and soft cheese. It can also be transmitted through direct contact with infected animals and animal health workers [2–4]. Brucellosis has relatively nonspecific symptoms, making it difficult to diagnose and differentiate from diseases like malaria, typhoid, or tuberculosis [4,5]. However, specific complications may occur, such as endocarditis, epididymo-orchitis and meningoencephalitis [6]. In addition to its clinical impact, brucellosis also has significant economic ramifications due to losses in animal production and time lost from work by patients [6,7].

Brucellosis has been prioritized in animal health for decades, with examples of successful eradication campaigns in cattle in Europe, the Americas and other high income countries [8– 10]. These programs have been shown to be successful technically and economically [8–10] yet there have been difficulties replicating these successes in other species. Despite the recognition of its importance by animal health systems, the high level of *Brucella* infections in many areas of the world in both human and animal hosts and the status as a neglected zoonosis by the World Health Organization (WHO), brucellosis is rarely prioritized by human health systems. [11,12]. To address this gap, researchers and public health institutes worldwide have initiated burden of disease assessments in humans based on the DisabilityAdjusted Life Year (DALY), a summary measure of public health widely used to quantify and compare disease burden. DALYs integrate the effects of mortality (Years of Life Lost, YLL) and morbidity (Years Lived with Disability, YLD) into a single metric and allow for the incorporation of different health states, as defined by the disease model [13]. In spite of the role that DALYs and other health metrics have played in informing national and global priorities, brucellosis DALYs estimates remain globally incomplete and infrequently undertaken. For instance, according to the WHO Foodborne Disease Burden Epidemiology Reference Group (FERG), brucellosis caused 393,239 (UI 143,815 – 9,099,394) foodborne illnesses in 2010, resulting in 124,884 (43,153 – 2,910,416) DALYs worldwide [14]. To put these results into context, FERG estimated that in 2010, a total of 33 (UI 25 – 46) million DALYs were caused by the foodborne diseases investigated [15]. These findings do not include the burden of direct transmission between animals and humans, thus, they are likely underestimated. Currently, the Global Burden of Disease (GBD) does not include brucellosis as a standalone cause of death and disability in their studies [16].

DALY calculations entail several methodological decisions and assumptions, which may limit comparability and interpretation of results generated by different studies, thus jeopardizing their relevance to policymakers as a prioritisation tool. Different burden estimates not only vary in terms of the geographic scope and temporal range they are conducted for, but also in fundamental methodological decisions and assumptions involved in the underlying calculations. Understanding both is critical to appreciating differences between studies and estimates, and a necessary part of the ongoing improvement in estimation efforts. This review will inspect key methodological choices underpinning DALY calculations. For instance, DALYs may be calculated from an incidence or a prevalence approach. The former approach considers the future health impact due to current exposures (YLD = incidence * duration * disability weight), while the latter approach considers the current health impact due to past exposures and emphasizes its contemporary impact on health systems (YLD = prevalence * disability weight) [17,18]. In both perspectives, disability weights (DWs) are an essential component. They translate morbidity into healthy life years lost, thus enabling comparison of morbidity and mortality; in other words, they estimate the disability scaled from 0 (perfect health) to one (death) [19]. Another crucial choice is the life expectancy table used for calculating YLLs, which defines the weight given to age-specific deaths. DALY calculations may also apply social weighting to give a different value to healthy life years lost at different ages (i.e., age weighting [19,20]) or time periods (i.e., time discounting [13]). Age weighting reflects the perspective that a year of life lived at one age is worth more than another. Similarly, time discounting indicates that the benefits and costs of today are valued more than those in the future. A final methodological choice is the construction of the disease model, which defines the health states that will be considered in the estimates and the possible transition between states [17]. In addition to these intrinsic methodological choices, DALY calculations are often hindered by data gaps and uncertainties, and different methods exist to deal with these limitations.

Dean et al. conducted two systematic reviews on specific brucellosis outcomes, namely its prevalence and clinical manifestation [4,21]; however, no comprehensive review on burden assessments has been carried out yet. Thus, this systematic review aims to identify existing brucellosis burden studies and to analyze their methodological choices, assumptions, and uncertainties. As a result of this assessment, knowledge gaps will be identified which need to be addressed to generate improved and comparable brucellosis burden of disease estimates.

The paper will focus on the human health burdens of brucellosis, it will not address the estimations of burdens in the livestock sector, which will be covered in other areas of the Global Burden of Animal Diseases programme (GBADs) [22] . Its contribution to GBADs is to improve understanding of the impact of animal diseases on society, including human health.

Zoonotic diseases that cause health issues and burdens to people and the animals they keep are problematic in society, identifying where burdens occur will help policy makers in their decisions on resource allocation and legislation.

## Methods

### Data sources and search strategy

This systematic literature review followed the Preferred Reporting Items for Systematic Reviews and Meta-analyses (PRISMA) Statement (S1 PRISMA) [23]. The protocol of this review was not registered. The databases PubMed, Embase and Web of Science were systematically searched, using terms covering DALY calculation and brucellosis. The search strategy, including Boolean operators, is presented in S2 Search strategy. A grey literature search was also conducted using a selection of databases (i.e., OpenGrey, OAIster, and the Journals Online project (JOLs)). A further search within the reference lists of included publications was performed to identify eligible burden studies which were not flagged by the initial search.

### Eligibility criteria

Peer-reviewed articles and grey literature published between January 1990 and July 2021 were included. Studies published before 1990 were omitted since the DALY metric was introduced in the early 1990s [13]. All studies quantifying DALYs were included, whether the primary purpose was a burden of disease assessment or not (e.g., cost-effectiveness studies). No language restrictions were applied. If necessary, help with translation was asked from other members of the Global Burden of Animal Diseases. Lastly, conference proceedings, abstracts, letters to editors and general correspondence were excluded.

### Data screening, selection, and extraction

Rayyan.ai [24] was used to manage citations and detect duplicates. First, duplicate entries were identified by considering the authors, the year of publication, the title of the article and the volume, issue, and page numbers. When in doubt, the abstract texts were compared. Subsequently, the sources were screened for eligibility (title – first step; abstract – second step; full-text – third step). The screening process was conducted by one researcher (CDB); when in doubt, articles were discussed with the study supervisor (BD). Finally, a database of all retrieved publications that met the eligibility criteria was created. For each eligible paper, an extraction grid (S3 extraction grid and definition) was used to gather data regarding study information and characteristics, i.e., data sources for epidemiological data, data adjustment, methodological perspective used to calculate YLDs and YLLs, and the DALYs result. The systematic review of Haagsma et al. [20] was used as a blueprint when creating the data extraction grid; in particular, the same categories proposed by Haagsma et al. [20] were used when collecting incidence and prevalence data. A separate grid was created (S3 extraction grid and definition) to collect the disease model information, i.e., the included health states and their transition probabilities, disease weights and durations. Additionally, a narrative summary for each study was made, including limitations and reasoning for the methodological choices made by the authors. Data extraction was performed by CDB, while NV reviewed.

## Results

### Flow of selected studies

Figure 1 shows a detailed flow diagram for selecting articles included in the systematic review. In total, 309 articles were retrieved. After the removal of duplicates and the screening for eligibility, 21 studies were retained for full-text screening. Two additional suitable studies were found through reviewing the list of references. In the end, thirteen studies were included for data extraction.

**Figure 1.**
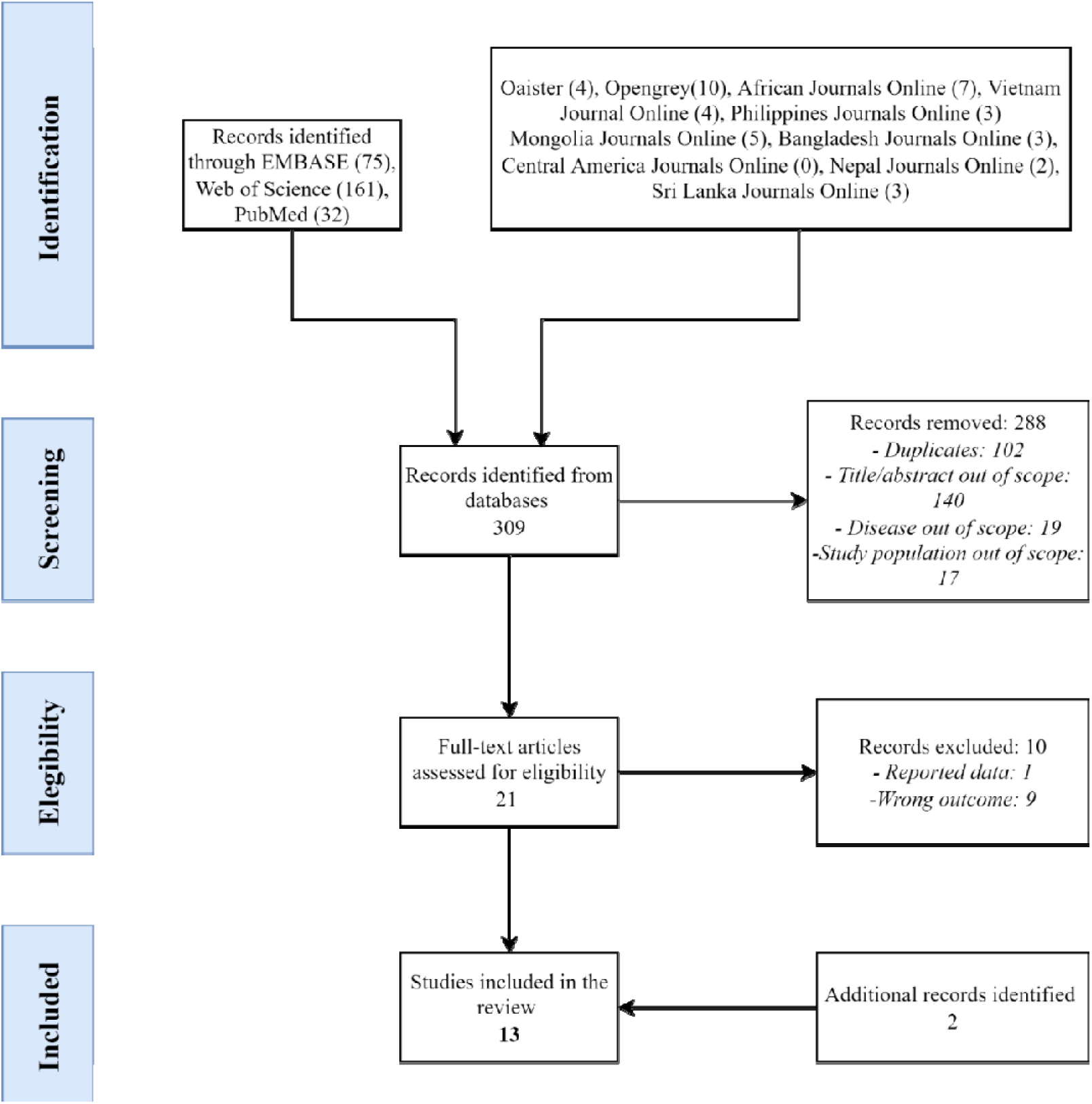
Flow chart of selected studies.

### Number of burden of disease studies per location and year of publication

Out of the thirteen eligible articles, only one estimated the brucellosis burden globally, the FERG study, by Kirk et al. [14]. Others were national or subnational studies, set in Iran (2) [25,26], Kenya (2) [27,28], Greece [29], India [30], Kazakhstan [31], Kyrgyzstan [32], Mongolia [33], Sudan [34], Taiwan [35], and Tajikistan [36](1 each) (Figure 2). One of the studies set up in Kenya [28] and the study in Sudan [34] were sub-national studies, where they presented only sub-national estimates.

**Figure 2.**
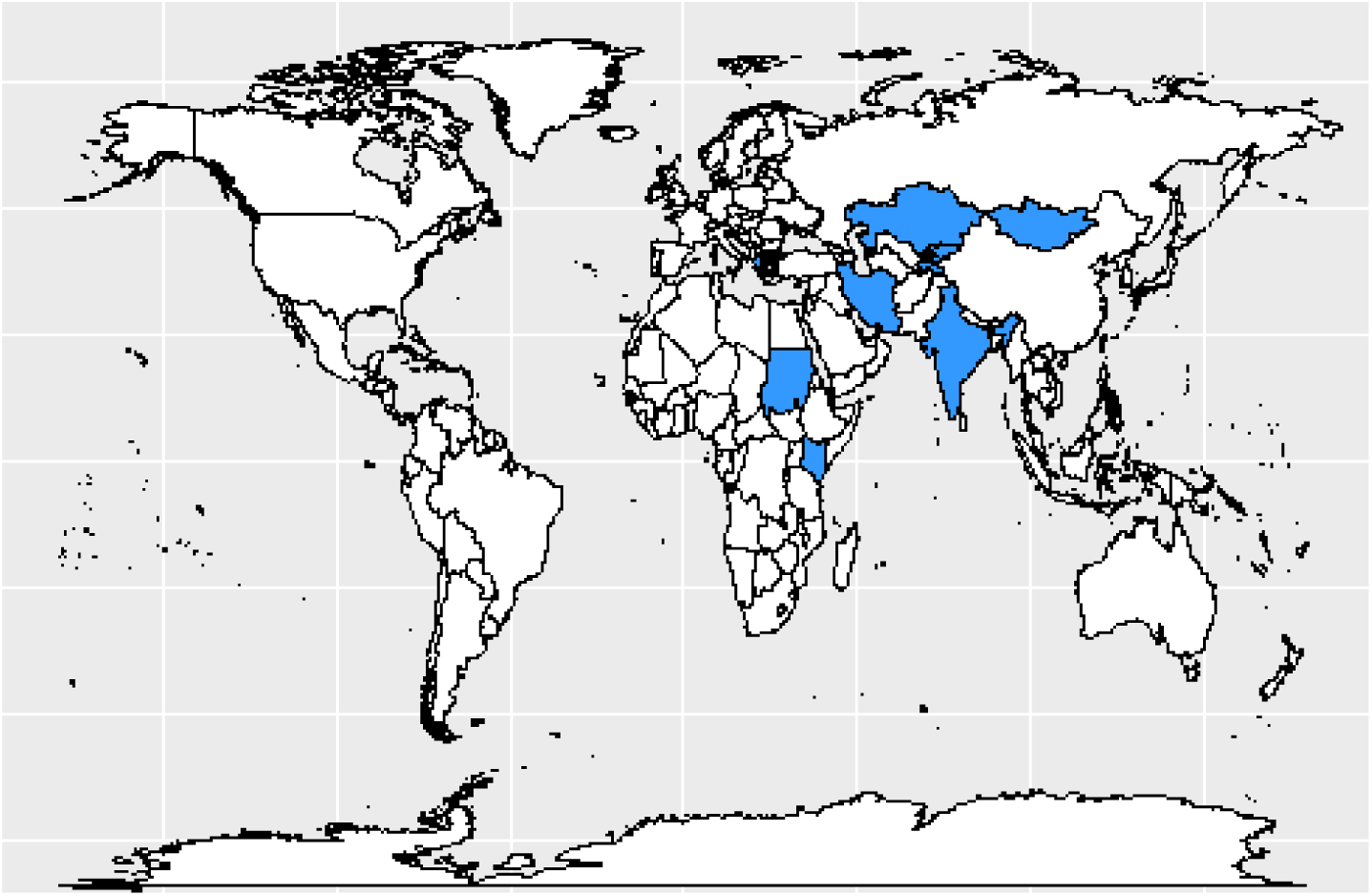
Countries with national or subnational *Brucella* burden studies estimates.

Regarding the publication years, the first burden study dates back to 2003, while the most recent one was published in 2021. In recent years, the number of published studies have increased (Figure 3).

**Figure 3.**
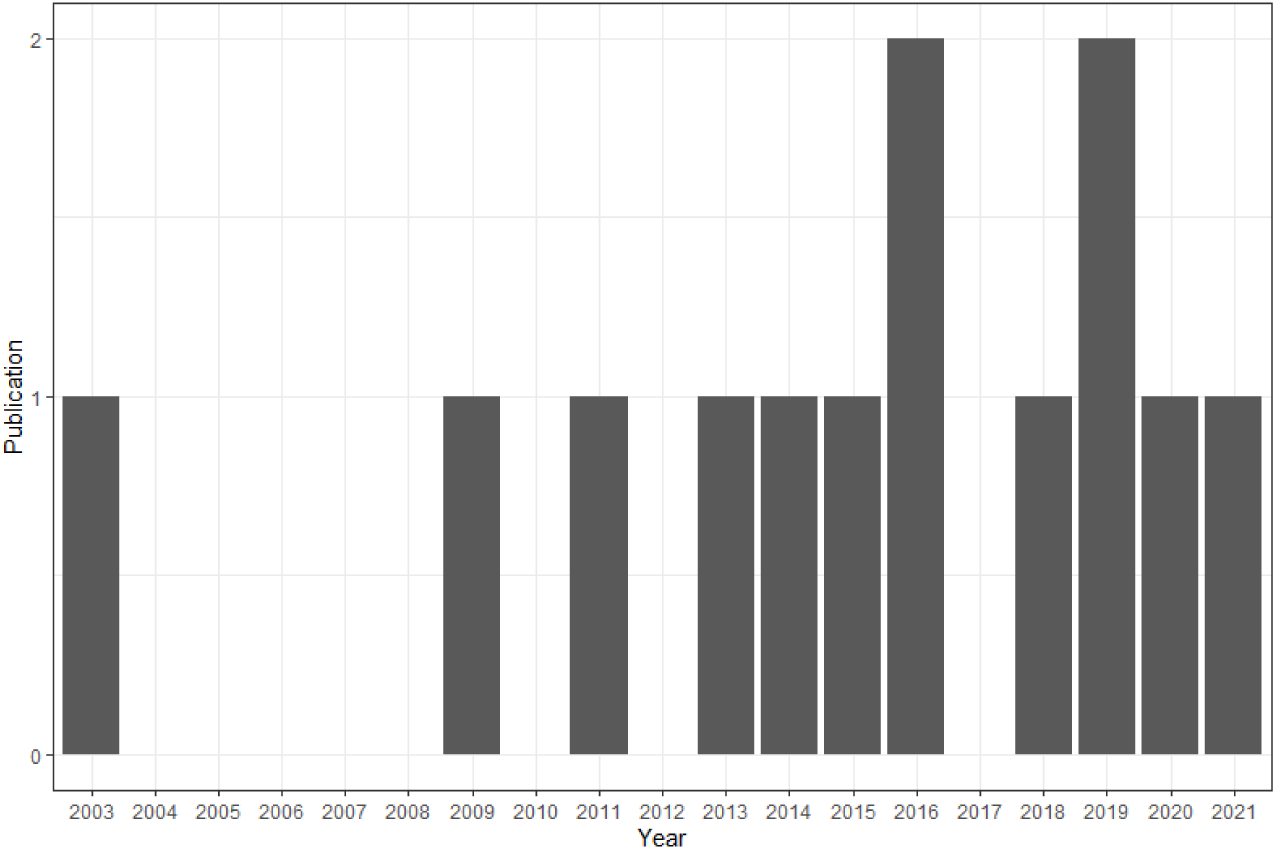
Existing *Brucella* burden of disease studies by year of publication.

### Epidemiological data

The selected studies used different epidemiological inputs as a source for DALYs calculation. Most of the studies, ten out of thirteen, retrieved incidence data from laboratory-confirmed cases [7,25,27–32,34,35]. In addition, three studies collected the information through previous literature [14,30,32] and one study used data from syndromic surveillance [26]. Lastly, one study [30] obtained the incidence by dividing the number of seropositive cases by the duration of seropositivity (estimated to be 10.9 years). Notably, studies from Counotte et al. [32] and Singh et al. [30] sourced data from multiple inputs, such as routine data and surveillance systems, and previous literature. The former employed different sources to validate the findings, while the latter analyzed particular subgroups [30,32]. Eight studies assessed mortality. Five studies sourced the mortality indirectly using case fatality rates based on previous literature [7,14,29,31,32]; for example, Charypkhan [31] used the case fatality reported by a German study, which used a German data set [37]. The remainder derived mortality data directly from the national surveillance system as in the case of Piroozi et al., or death registry as in the case of Navaghi et al. and Lai et al. [25,26,35].

#### Data adjustments

Four studies considered and addressed the underestimation of incidence using two different approaches. Three studies used a data-driven approach. The study conducted in Kyrgyzstan [32] used prevalence data to address the underestimation in the official data. The study of Kirk et al. [14] computed a multiplier by comparing human incidence data from the World Organization for Animal Health (OIE) on countries “free of brucellosis in livestock” and the literature review by Dean et al. [21]. In contrast, Gkogka et al. [29] computed a multiplier from previous literature [38,39]. The other method to address underestimation was based on expert opinion. The value of the multiplier in the study by Piroozi et al. [26] was established by an expert panel. Interestingly, Brink conducted a scenario analysis with different underestimation rates, but the main results presented in the study did not consider underestimation [27].

Three studies assessed the possibility of missing values among model parameters and handled them in various ways. Specifically, Counotte et al. [32] used data from neighboring countries or overlapping regions for missing prevalence data; Kirk et al. [14] dealt with missing incidence data by computing the Bayesian log-normal random effect. Finally, Singh et al. carried out data triangulation to estimate the missing value of specific subgroups, such as the number of para-veterinarians [30].

### Methodological choices

#### General disability-adjusted life years methodology

Not all studies considered YLLs caused by brucellosis. In fact, five studies explicitly stated that they did not consider them [25,27,28,30,34], while one study did not provide any information (Table 1) [36]. In the study of Naghavi et al. [25], the official registry did not report mortality data for brucellosis. Brink, Singh et al. and Alumasa et al. [27,28,30] explained their decision on the basis of previous studies and national surveys that did not report brucellosis mortality data. Alternatively, Tamador did not estimate YLLs from its perspective that “brucellosis is not a fatal disease” [34]. The remaining studies (N = 7) employed life expectancy tables from different sources to calculate YLLs (Table 1). Three studies [14,31,32] used WHO 2050 [40]; three studies used country-specific life tables [26,29,35]; whilst one study did not specify the life table chosen [7]. Finally, all the studies (N=13) followed an incidence-based approach to calculate YLDs. No study adjusted for comorbidity.

**Table 1.** Summary of disability-adjusted life year calculation methods for brucellosis

| Region | Study | General |  | YLL | YLD |  |  |  | Results |  |
| --- | --- | --- | --- | --- | --- | --- | --- | --- | --- | --- |
|  |  | Discount rate | Age weighting | Life table | Approach | Duration source | Disability weights source | Archetypes used in the disease model | Incidence | DALYs per CASE |
| Global | Kirk et al., 2015 | No | No | WHO 2050 | Incidence | Literature (Dean et al., 2012) | GBD 2010 | Disease model 3 | 832,633 | 0.3 |
| National | Counotte et al., 2016 | No | No | WHO 2050 | Incidence | Literature (Kirk et al., 2015) | GBD 2010 | Disease model 3 | 2,435 | 0.29 |
|  | Gkogka et al., 2011 | No | No | Country-specific | Incidence | Literature (Roth et al., 2003) | GBD 1990 | Disease model 2 | 79 | 0.62 |
|  | Roth et al., 2003 | Yes | Yes | Not stated | Incidence | Literature (Beklemishev, 1964) | GBD 1990 | Disease model 2 | 1,482 | 0.62 |
|  | Piroozi et al., 2019 (*for incidence data S3*) | Yes | No | Country-specific | Incidence | Expert | Burden of disease and injury in Iran 2003 | Disease model 2 |  | 0.17 |
|  | Singh et al., 2018 | No | No | - | Incidence | Literature (Roth et al., 2003) | GBD 1990 | Disease model 1 | - | 0.90 |
|  | Charypkhan et al., 2019 | No | No | WHO 2050 | Incidence | Literature (Bosilkovski et al., 2010) | Dean et al. (chronic brucellosis) | Disease model 2 | 1,334 | 0.50 |
|  | Kurbonov et al., 2016 | Not stated | Not stated | - | Incidence | - | - | - | - | 1.69 |
|  | Alumasa et al., 2021 | No | No | - | Incidence | Literature (Roth et al., 2003) | Dean et al. (acute brucellosis) | Disease model 1 | - | 0.81 |
|  | Tamador, 2014 | Yes | Yes | - | Incidence | Not stated | GBD 1990 | Disease model 1 | 176 | 1.00 |
|  | Brink, 2013 | Yes | No | - | Incidence | Literature (Kunda et al., 2007) | Dean et al. (acute brucellosis) | Disease model 1 | 75,256 | 0.10 |
|  | Lai et al., 2020 | No | No | Country-specific | Incidence | Literature (Kirk et al., 2015) | WHO/FERG | Disease model 2 | - | - |
|  | Naghavi et al., 2009 | Yes | Yes | - | Incidence | Expert | Expert panel | Disease model 1 | - | - |

Five out of the thirteen studies applied age-weighting or time discounting rates. In particular, three studies applied both age-weighting and time discounting [7,25,34] (table 1). Tamador and Naghavi et al. chose, respectively, a rate of 0.04 and 0.3 [25,34]. In Roth et al.’s study, different time discounting rates (3% and 5%) were applied, with an age-weighting of 0.16 [7]. Piroozi et al. and Brink only considered time discounted at 3% [26,27].

Table 1 shows the Different DALYs per case across the studies. There is great variation in the value of DALY per case; the minimum DALY per case is 0.10 presented by Brink [27], followed by the value of Piroozi et al. [26] (0.17). On the other end of the spectrum, there are the estimates presented by Kurbonov et al. [36] and Tamador [34], respectively 1.69 and 1.00.

#### Disease model choices

##### Health states

Out of the thirteen studies that calculated DALYs for brucellosis, twelve provided information regarding the disease model, including the different health states. All the studies applied one of the three archetypes, which are outlined in Figure 4. Kirk et al. and Counotte [14,32] used the same disease model (Figure 4, Brucellosis disease model 3), developed by Kirk et al., entailing six different health states (acute, severe, moderate, chronic, orchitis and death). Furthermore, they defined the probability of developing each health state. Kirk et al. [14] reported the different GBD proxy health states used to determine the disability weights that they implemented. Additional details can be found in the extraction grid (S3 Extraction grid and definitions). Although Counotte [32] employed Kirk’s health model, some differences are identifiable, especially in the duration of chronic brucellosis, but no explanation is provided for these differences. Five studies presented two health states (a general “brucellosis” and death) (Figure 4, Brucellosis disease model 2). The rest of the studies (N=5) reported only brucellosis as a health state (Figure 4, Brucellosis disease model 1). They did not report death as a health state, because they did not calculate YLLs, or did not consider brucellosis as a fatal disease.

**Figure 4.**
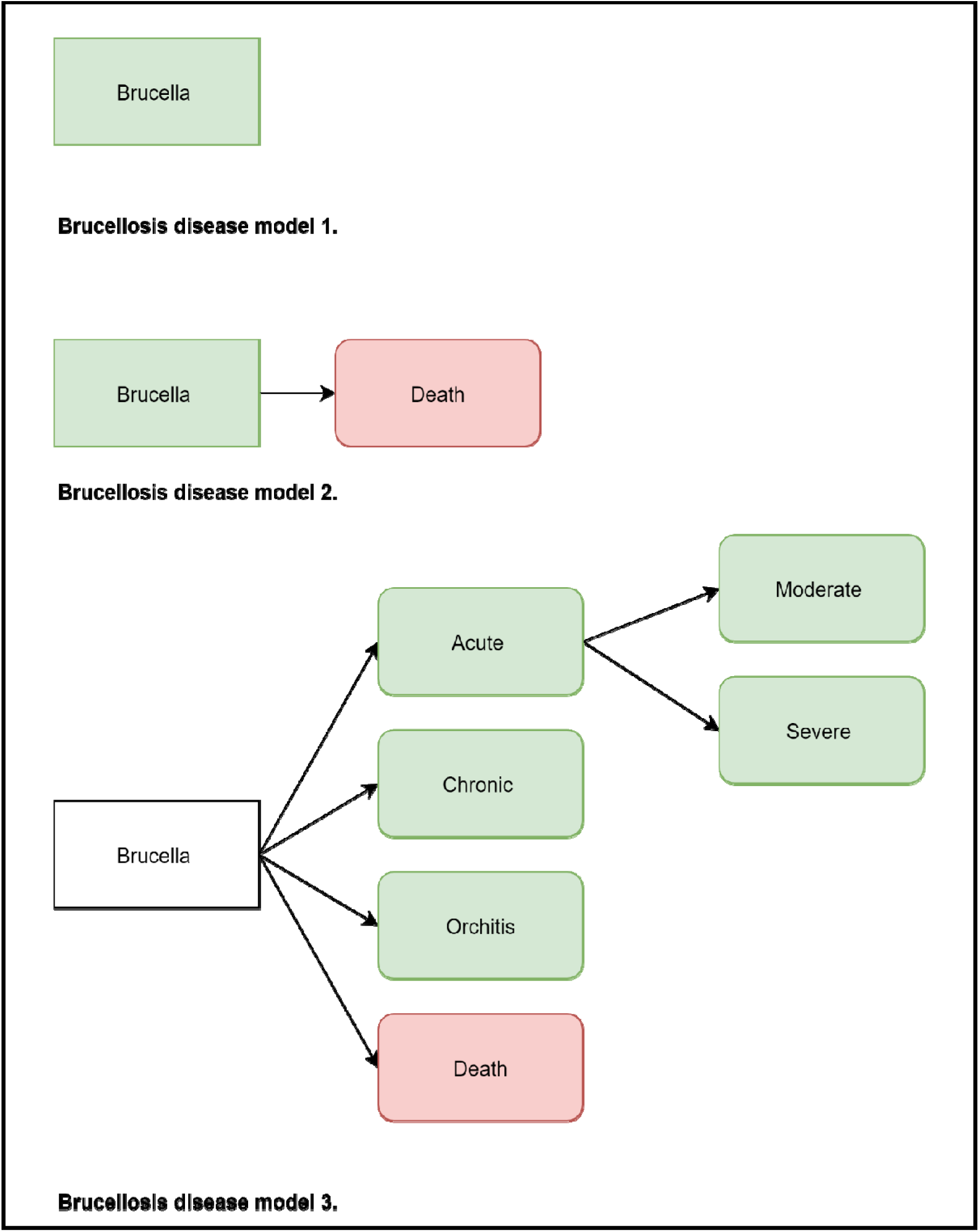
Disease models for brucellosis. Rectangles define the parent nodes, while rounded rectangles define the child nodes. White nodes do not contribute directly to the DALY estimate; green nodes contribute YLDs; and red nodes contribute YLLs. These models are not based on the biological or clinical pathway but they are computational disease models.

##### Duration

The burden assessments retrieved the disease duration from different sources (Table 1). Whilst eleven studies reported how they obtained it, two studies did not specify the process [25,36]. Nine studies derived the duration of brucellosis from the literature. Among those, three studies [28–30] retrieved this parameter from Roth et al. [7]. Roth et al. initially assumed 4.5 years, based on Beklemishew [41], but for the nature of the study, the median of the cumulated discounted DALYs was assumed, corresponding to 3.11 years. For this reason, discrepancies could be found in the studies that retrieved duration from Roth et al. [7]. While Gkogka et al. used the value of 3.11 years, Singh et al. and Alumasa et al. kept 4.5 years as the duration of brucellosis. Kirk et al. used the result provided by the literature review conducted by Dean et al. [4] and differentiated between acute infections (14 days) and chronic infections (6 months). Counotte et al. [32] employed the data from Kirk et al. as the source for the disease duration of brucellosis [14]. The burden estimated by Lai et al. [35] also cited the duration presented by Kirk et al. [14], however, they did not regard the different health states separately but instead assumed an overall disease duration of 0.038 years (two weeks). Charypkhan [31] employed the duration from Bosilovski et al. [42] (0.21 years or ten weeks) and confirmed it with the data reported by Dean and colleagues [21]. Brink derived it (0.5 years or six months) from studies by Kunda et al., Kursun et al. and Muriuki et al. [43–45] (S3 extraction grid).

Two studies, both set in Iran, used expert opinions to estimate the duration of brucellosis. Piroozi et al. described that they held an expert panel with infectious diseases specialists [26]. The duration has been assumed to be 0.75 years. For *The burden of disease and Injury in Iran 2003*, the duration was estimated with expert input from Iranian disease epidemiologists and clinical specialists [25]. The resulting disease duration was not reported. The study by Tamador and colleagues assumed three years for the duration of illness; however, it failed to report the source [34]. More information can be found in Table 1 and in S3 extraction grid.

##### Disability weights

Another sensitive methodological choice that needs to be considered is the disability weight (DW). Out of thirteen studies, twelve reported the source for their DW (Table 1). It varied from 0.053 [14] to 0.23 [25,26]. Two studies [14,32] derived the DW from proxy health indicators in the GBD 2010 [46]. For example, their DW for acute and severe brucellosis corresponded to the GBD 2010 definition of “infectious disease: acute episode, severe” (S3 extraction grid and definitions). Lai et al. [35] retrieved the DW (0.092) from Kirk et al., [14] and have subsequently calculated the weighted average between acute episode severe (DW=0.133) and acute episode moderate (DW=0.051). Alternatively, four studies [7,29,30,34] used DWs from GBD 1990 [13]. Studies that referred to the GBD 1990 have a DW of 0.2, also defined as a disability of class II. A DW of 0.2 according to the GBD 1990 correspond to a condition that is “painful and affecting occupational ability even during period of remission” [13]. Furthermore, three studies retrieved the DW from Dean et al. [4]. Dean et al.[4] proposed two different DWs, based on GBD 2004: 0.15 for chronic brucellosis and 0.19 for acute brucellosis. On the one hand, Charypkhan et al. [31] utilized 0.15, which is the DW proposed by Dean et al. for chronic brucellosis [4]. On the other hand, Brink [27] took the acute state (0.19) as reference. Alumasa et al. [28] also refer to Dean et al. [4] as the source for deriving their DW of 0.18. Piroozi et al. (0.23) sourced the DW from the *Burden of disease and injury in Iran in 2003 using the Delphi method* [25]. A summary can be found in Table 1.

#### Source attribution

Four studies explored and took into account different routes of transmission for brucellosis (S3 Extraction grid). Three studies considered transmission via food as a transmission route. They retrieved the percentage attributable to foodborne exposure from previous literature [29,35] or from global expert elicitation [14]. In contrast, the forth study [30] considered occupation as a transmission route. Information was retrieved from data regarding the prevalence of human brucellosis among veterinary personnel, abattoir workers and livestock farmers from the published scientific literature. More information on the attribution source can be found on S3 extraction grid and definition.

### Uncertainty analyses: Analyzing model specification, parameters and uncertainty

Four studies conducted their own scenario analysis. Different elements were used when doing a scenario analysis: different life expectancy tables (country-based and West level 26) [29], discount rates (0.3 and 0.5) [7], with or without age weighting and time discounting [30], and different degrees of underestimation (0, 5, 20, 50, 75, 90%) [27]. Uncertainty was modelled by five studies using Monte Carlo simulations [7,14,29,30,32].

## Discussion

This review provides a comprehensive overview of the number and types of studies that calculated the burden for human brucellosis that were published from January 1990 to July 2021. The aim was to identify existing brucellosis burden studies, and analyze their methodological choices, assumptions, and uncertainties. In total, thirteen studies met the inclusion criteria. The OIE World Animal Health Information System database collects information on animal health around the world. According to the OIE data from 2005 to 2022, *Brucella* is reported in animals in the Mediterranean region, in the Middle East, Russia, Sub-Saharan Africa, in some countries of Latin America and in central and south Asia [47]. Due to brucellosis’ mode of transmission, it can be assumed that it affects humans in the same locations or in people who travel to these areas. Despite repeated documentation of brucellosis within animal hosts, there is no human burden data available in some of the locations that are affected. Indeed, most of the studies included in this review were from Eastern Europe, Asia, and Sub-Saharan Africa. No studies were identified from North, Central and South America or

South-East Asia. This is in line with the observations by Dean et al., who noted in their reviews [4,21] that the same geographical areas were missing brucellosis burden data. This could potentially reflect either a lower disease presence or poorer brucellosis surveillance. Thus, the (national) studies that we found present only a limited picture of the burden of brucellosis.

Our review revealed large methodological variations in available burden estimates for brucellosis. Indeed, differences are visible when scrutinizing the DALYs per case presented by the articles (Table 1). The variations in the DALYs per case highlight the impact of the methodological choices and assumptions proposed by the authors (Table 1). For instance, Brink et al. [27] have the lowest DALYs per case (0.10); this is due to methodological choices, such as not accounting for mortality, applying time discounting, and having chosen a shorter duration (6 months), and a DW of 0.19. In contrast, the study conducted by Gkogka et al. [29] presented different DALYs per case based on different methodological choices. Gkogka et al. accounted for YLL, with a case fatality of 2%. Time discounting and age-weighting were not applied. Additionally, the chosen duration and DW were 3.11 years and 0.2, respectively. These two examples illustrate the considerable direct impact of the methodological choices on the estimates. This implies that comparisons of available (national) estimates with other countries should only be done with caution. A method to enhance direct comparison between the different DALYs estimates would be to recalculate the DALYs using a single consistent methodology or by choosing one DALYs per case presented by one of the studies and applying it across the studies.

In what follows, we describe the main implications of the methodological differences observed. All the studies employed an incidence-based approach. The incidence-based approach reflects the future burden based on current events, where all health outcomes are assigned to the initial event [20]. For the burden of brucellosis (an infectious and foodborne disease), the incidence-based approach is often mentioned to be preferable because it is sensitive to current epidemiological trends and it is more consistent with YLLs, which by definition follow an incidence approach (mortality can be seen as the incidence of death) [17,18]. Additionally, laboratory-confirmed cases were often used to compute the incidence. However, reliance on solely laboratory-confirmed cases would result in underestimation [20]. A correction for potential underestimation was only included in four studies. Underestimation of the brucellosis burden has implications in priorities settings and could undermine the value of the studies as advocacy tool.

The majority of mortality data were obtained indirectly. Moreover, some studies stated that the official registries reported no data on mortality. This could happen due to a lack of data, indicating that fatal outcomes of brucellosis are rare, or due to inadequate cause of death reporting systems. Calculating disease burden estimates requires high-quality mortality data. The absence of those has an important consequence in the calculation of YLLs, although in the case of brucellosis, most of the burden relies on YLDs. Nevertheless, the omission of YLLs will cause an underestimation of the burden.

Differences in the duration values were observed, which revealed that there is uncertainty surrounding the length of this disease. The duration utilized by the articles varied from two weeks to 4.5 years. Counterintuitively, the duration did not have a connection with the archetype (see Figure 4) that has been used. For example, the duration of 4 or 4.5 years was used for the estimates of Singh et al. and Roth et al. and they applied the first and second archetype of brucellosis’ disease model respectively. Meanwhile Kirk et al. and Counette et al. applied archetype 3 but they used shorter durations also for the chronic phase (six months).

All studies took the duration data from previous literature or expert opinion, and no study used duration that was directly data-driven. In particular, studies that retrieved it from previous literature used the same few sources, which could probably be context-specific and/or outdated. For instance, as in the case of Beklemishew [41], used by Roth et al., the study was based on the clinical data of 1000 untreated patients in the Russian Federation. This raises questions on the validity of the selected values in contemporary contexts. Another finding relating to the disease model was that most studies did not consider different health states and symptoms. This could be caused not only by the lack of clinical data on brucellosis sequelae but also by the lack of knowledge on the disease course. Additionally, none of the studies referred to abortions in pregnant women as a health state, even though different studies provide evidences of the adverse effects of brucellosis on pregnancy outcome [48–50]. Few studies took into account the routes of transmission of brucellosis and the relative importance of these routes. A deeper understanding of the transmission could help gain a better familiarity with sources of attribution and help develop effective prevention measures. Finally, we observed that not many disease burden studies assessed uncertainty. Ideally, studies should clearly describe the uncertainties in their data and methods, and quantify them. Indeed, this will strengthen the validity of the results and allow better comparability across different studies.

### Limitations

The review has some limitations. First, ongoing estimates or studies that were performed but not documented in peer-reviewed articles or grey literature could potentially be missing. To limit this possibility, reference lists were checked. Second, the search strategy was only conducted in English. This could potentially lead to missing out on some eligible and valid studies in other languages. Finally, there is no standardized way of evaluating DALYs calculations or disease models currently in place. Thus for this review, we had to come up with an approach ourselves, which was inspired by previous literature along with the work done by the European Burden of Disease Network [51].

### Future prospects

This review aimed to report different methodological choices that have been made in assessing the burden of brucellosis in humans. The findings of the review can serve as a base to improve future burden estimates, especially global analyses such as FERG and GBD. The OIE data [47] showed that brucellosis is spread worldwide; nevertheless, human burden estimates are only available for a limited number of countries where animal brucellosis is reported to occur. Better synergies between animal and human data for brucellosis will not only provide better estimates but also a clearer view on transmission and on effective interventions. Indeed, this review will support the GBADs programme to develop a systematic methodology to describe the impact of animal diseases on society, including human health, and to close the gap between human and animal health.

From the review, it appears that incidence data, and especially mortality and duration, should be better addressed with more data-driven approaches. Consequently, further research needs to be done for these parameters across multiple countries. Better routine reporting, new clinical studies focusing on duration and incidence, and better mortality registries would help fill these gaps. Additionally, more evidence is needed on the link of specific brucellosis symptoms with health states to better understand the course of the disease. Finally, exploring sources of attribution will give a better understanding of the transmission and prevention of brucellosis.

This review found a great variety in the methodological choices and assumptions used to calculate DALYs and numerous inconsistencies in reporting methods and assumptions. Thus, there is a need for a more standardized reporting system for DALYs estimates, which could resemble a checklist that reports the methodological choices and assumptions. The adoption of this tool will enhance the transparency and understanding of the methodological choices and the subsequent reuse of these estimates for prioritisation purposes.

## Conclusion

This review aimed to explore the different methodological choices and assumptions used to quantify the DALYs for brucellosis. The results suggested that some parameters carry considerable uncertainty, particularly mortality and disease duration. This highlights the importance of strengthening routine reporting, collecting better mortality data and conducting further research on the course of brucellosis. Additionally, estimates of DALYs will benefit from a deeper understanding of the symptoms of the disease and the different sources of attribution. Current reporting of methodological choices should be improved to enhance transparency, comparability, and consistency of brucellosis burden estimates. Finally, within the scope of the GBADs programme, this review supports the development of a systematic methodology to describe the impact of animal diseases on society, including human health, and to strengthen the connection between human and animal health.

## Supporting information

S1 PRISMA checklist

S2 search strategy

S3 extraction grid

## Data Availability

All data produced in the present work are contained in the supporting documents

## Supporting Information

S1 PRISMA Checklist S2 Search Strategy

S3 Extraction Grid

## Acknowledgements

This work was supported, in whole or in part, by the Bill & Melinda Gates Foundation INV-005366. Under the grant conditions of the Foundation, a Creative Commons Attribution 4.0 Generic License has already been assigned to the Author Accepted Manuscript version that might arise from this submission.

## Declarations

This research is on behalf of the Global Burden of Animal Diseases (GBADs) programme which is led by the University of Liverpool and the World Organization for Animal Health (OIE) (https://animalhealthmetrics.org/). This research is supported through the Grant Agreement Investment ID INV-005366 with the Bill & Melinda Gates Foundation and the UK Foreign, Commonwealth and Development Office (FCDO). GBADs case studies receive additional funding from the following: European Commission, Australian Centre for International Agricultural Research (ACIAR), Brooke Foundation and the Food and Agriculture Organization of the United Nations (FAO). A full list of the GBADs collaborators can be accessed here: https://animalhealthmetrics.org/acknowledgements.

