## Supplementary material for "Methodological choices in brucellosis burden of disease assessments: A systematic review": S2 search strategy

The search was carried out on the 23 July 2021.

### Embase

1 Brucella/ 4376
2 brucellosis/ 11075
3 disease burden/ 26005
4 disability-adjusted life year/ 2665
5 years of life lost.mp. 2326
6 years lived with disability.mp. 970
7 Disability-adjusted life years.mp. or disability-adjusted life year/ 5181
8 cost-effectiveness.mp. or "cost effectiveness analysis"/ 186962
9 1 or 2 13341
10 3 or 4 or 5 or 6 or 7 or 8 217460
11 9 and 10 75

### History Web of Science

16
**(#14) AND #13** and **Humans** (Search within all fields)
Edit
Add to Search
45
15
**(#14) AND #13**
Edit
Add to Search
161
14
**((((((((#12) OR #11) OR #10) OR #9) OR #8) OR #7) OR #6) OR #5) OR #4) OR #3**
Edit
Add to Search
259,510
13
**(#1) OR #2**
Edit
Add to Search
12,407
12
**TS=(cost-effectiveness)**
Edit
Add to Search
108,176
11

**TS=(YLD)**
Edit
Add to Search
431
10
**TS=(YLL)**
Edit
Add to Search
568
9
**TS=(years lived with disability)**
Edit
Add to Search
10,904
8
**TS=(years of life lost)**
Edit
Add to Search
11,601
7
**TS=(DALYs)**
Edit
Add to Search
2,463
6
**TS=(DALY)**
Edit
Add to Search
3,105
5
**TS=(Disability-adjusted life years)**
Edit
Add to Search
3,924
4
**TS=(Disability-adjusted life year)**
Edit
Add to Search
3,924
3
**TS=(Burden of disease)**
Edit
Add to Search
134,584
2
**TS=(Brucella)**
Edit
Add to Search
9,294
1
**TS=(Brucellosis)**
Edit
Add to Search
7,672

#### PubMed

| Search number | Query |
|---|---|
| **15** | (((((((((Burden of disease[Title/Abstract]) OR (Disability-adjusted life year[Title/Abstract])) OR (Disability-adjusted life years[Title/Abstract])) OR (DALY[Title/Abstract])) OR (DALYs[Title/Abstract])) OR (years of life lost[Title/Abstract])) OR (years lived with disability[Title/Abstract])) OR (YLL[Title/Abstract])) OR (YLD[Title/Abstract])) OR (cost-effectiveness[Title/Abstract])) AND ((brucellosis[Title/Abstract]) OR (brucella[Title/Abstract])) |
| **14** | ((((((((Burden of disease[Title/Abstract]) OR (Disability-adjusted life year[Title/Abstract])) OR (Disability-adjusted life years[Title/Abstract])) OR (DALY[Title/Abstract])) OR (DALYs[Title/Abstract])) OR (years of life lost[Title/Abstract])) OR (years lived with disability[Title/Abstract])) OR (YLL[Title/Abstract])) OR (YLD[Title/Abstract])) OR (cost-effectiveness[Title/Abstract]) |
| **13** | (brucellosis[Title/Abstract]) OR (brucella[Title/Abstract]) |
| **12** | cost-effectiveness[Title/Abstract] |
| **11** | YLD[Title/Abstract] |
| **10** | YLL[Title/Abstract] |
| **9** | years lived with disability[Title/Abstract] |
| **8** | years of life lost[Title/Abstract] |
| **7** | DALYs[Title/Abstract] |
| **6** | DALY[Title/Abstract] |
| **5** | Disability-adjusted life years[Title/Abstract] |
| **4** | Disability-adjusted life year[Title/Abstract] |
| **3** | Burden of disease[Title/Abstract] |
| **2** | brucella[Title/Abstract] |
| **1** | brucellosis[Title/Abstract] |
